# A comparison of four commercially available RNA extraction kits for wastewater surveillance of SARS-CoV-2 in a college population

**DOI:** 10.1101/2021.06.01.21257858

**Authors:** Megan O’Brien, Zachary C. Rundell, Michelle D. Nemec, Laura M. Langan, Jeffrey A. Back, Joaquin N. Lugo

## Abstract

Localized wastewater surveillance has allowed for public health officials to gain a broader understanding of SARS-CoV-2 viral prevalence in the community allowing public health officials time to prepare for impending outbreaks. Given variable levels of virus in the population through public health interventions, proper concentration and extraction of viral RNA is a key step in ensuring accurate detections. With many commercial RNA extraction kits and methodologies available, the performance of 4 different kits were evaluated for SARS-CoV-2 RNA detection in wastewater, specifically focusing on their applicability to lower population densities such as those at university campus dorms. Raw wastewater samples were collected at 4 sites on a college campus over a 24 hour period as a composite sample. Included in these sites was an isolation site that housed students that tested positive for Covid-19 via nasopharyngeal swabs. These samples were analyzed using the following kits: Qiagen All Prep PowerViral DNA/RNA kit, New England BioLabs Monarch RNA MiniPrep Kit, and Zymo Quick RNA-Viral Kit, and the Zymo Quick-RNA Fecal/Soil Microbe MicroPrep Kit. All four sites were processed according to the manufacturer’s guidelines. Extractions were then quantified with RT-qPCR one-step reactions using an N2 primer and a linearized plasmid standard. While the Zymo Quick-RNA Fecal/Soil Microbe MicroPrep Kit (also known as the Zymo Environ Water RNA Kit) only recovered approximately 73% (+/- 38%) SARS-CoV-2 RNA compared to the Zymo Quick-RNA Viral kit, it was the most time efficient kit to yield comparable results. This extraction kit had a cumulative processing time of approximately five hours compared, while the other three kits had processing times between approximately 9 and 9.5 hours. Based on the current research, the most effective kits for smaller population densities are pellet based and include a homogenization, inhibitor removal, and RNA preservation step.

**Highlights:**

- Samples from smaller population densities make concentrating RNA vital for detection
- Pelleting may provide for more timely concentration and extraction of SARS-CoV-2 RNA
- RNA shields and PCR inhibitor removal may increase detection of RNA during RT-qPCR

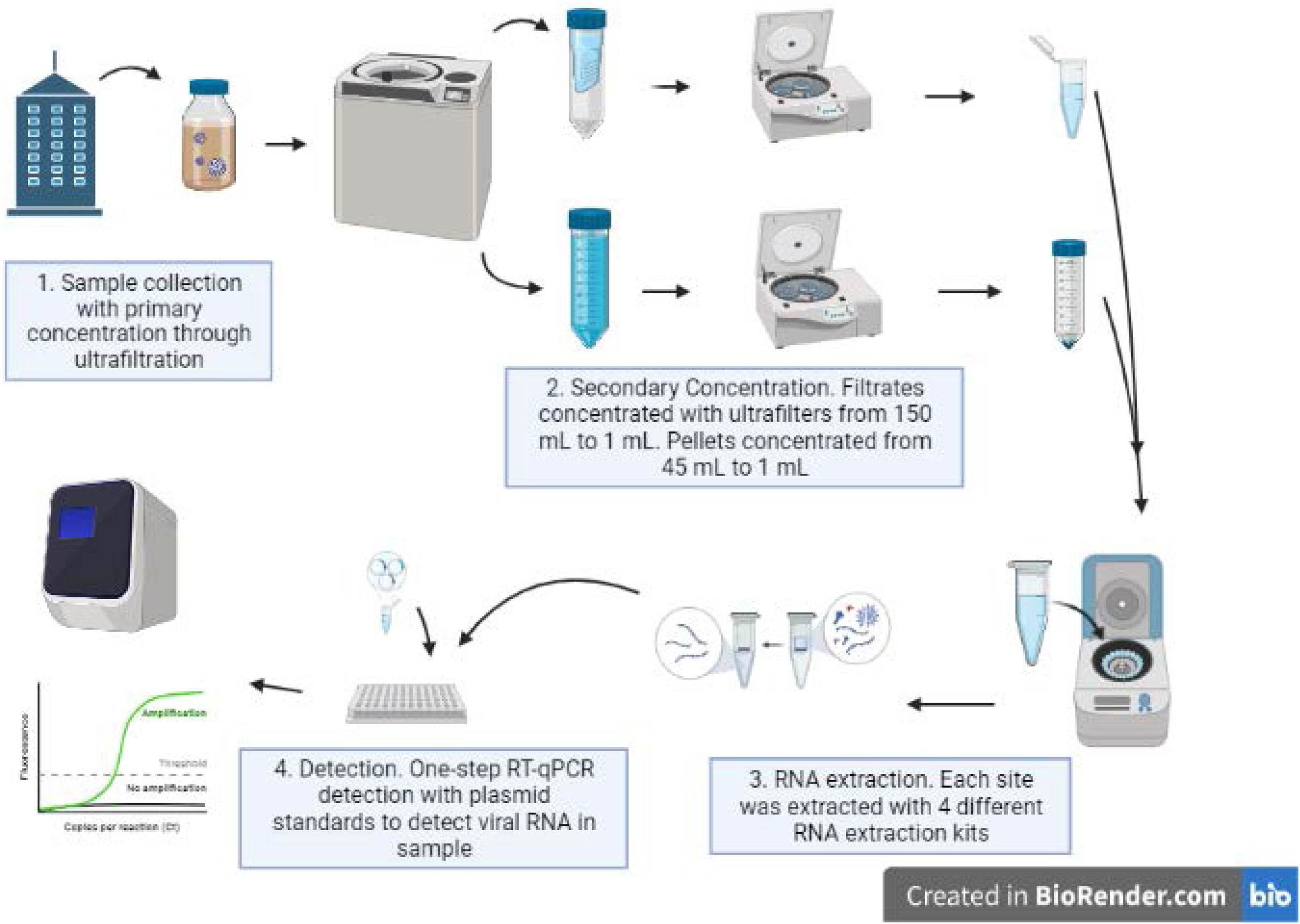

## 1. Introduction

On January 31, 2020, the WHO declared COVID-19 a Public Health Emergency of International Concern (Jee, 2020). Widespread testing of those with and without COVID-19 symptoms, in combination with the numerous vaccine roll outs is vital to curtailing the current pandemic and future outbreaks. The current estimate of infected individuals is believed to be underestimated worldwide, with numerous nations initially encouraging testing for those only with symptoms. Therefore, those who are presymptomatic or asymptomatic are often less likely to be identified, thus posing a significant potential for transmission, with studies estimating that asymptomatic or presymptomatic transmission could be responsible for up to 50% of new cases (Ghandi et. al 2020, Moghadas et. al 2020).

A variety of diagnostic testing methods are available to determine if individuals are infected with COVID-19. Current diagnostic testing involves the collection and PCR analysis of infected cells and bodily fluids for the SARS-CoV-2 virus by drawing blood or collecting samples from the nose, mouth, throat, or lungs (Ravi et. al 2020). While useful, these testing methods are hazardous, resource-intensive, and invasive (Binnicker 2020).

Wastewater based epidemiology offers a promising method of Covid-19 surveillance that may solve some of these pressing issues. Although a relatively new field, it has conventionally been successfully used to estimate use of legal and illegal drugs of abuse and to evaluate human exposure to contaminants and pathogens as summarized in Lorenzo and Picó (2019). Active monitoring of SARS-CoV-2 RNA in wastewater can be a useful tool for identifying hotspots and has been demonstrated to serve as an early warning system for new outbreaks (Xagoraraki and O’Brien, 2019, Venugopal et. al 2020, Betancourt et. al 2021). This method is unique as it allows researchers to survey large groups of people quickly with fewer resources and staff. Wastewater based epidemiology is also less intrusive compared to nasal swabs and reduces occupational exposure to SARS-CoV-2. Perhaps the largest benefit of wastewater surveillance is its efficiency and ability to view community prevalence. Thus, areas with higher viral copies may be focused on for individual testing efforts and public health interventions. Wastewater surveillance works due to SARS-CoV-2 RNA being detectable in the feces of both symptomatic and asymptomatic individuals (Hart and Halden, 2020, Mizumoto et. al 2020, Treibel et. al 2020), even after the individuals no longer had respiratory symptoms (Zheng et. al 2020, Mesoraca et. al 2020, Jones et. al 2020).

Wastewater based epidemiology is a powerful tool that can provide vital information about the spread of Covid-19 and can be useful in prioritizing diagnostic PCR testing. It has demonstrated its ability to effectively aide in detecting affected individuals so students could be tested or isolated to prevent further spread on a college campus (Betancourt et. al 2021). Unfortunately, a need exists for standardized techniques in applying this method to Covid-19 surveillance (Ahmed et. al 2020a). As populations contributing to the wastewater vary in their characteristics, so does the overall matrix of the wastewater sample (Kitajima et. al 2020). Consequently, numerous methods of sample processing exist, which can make it difficult for researchers to choose an optimal method for accurate detection. Furthermore, sampling in small population densities adds another layer of complexity to detection due to a potential for lower viral loads than typical in municipalities. Monitoring in communities with low incidence has previously demonstrated high PCR Ct values and hence variable or unquantifiable data being collected due to very low concentrations of the viral fragment in the collected samples (D’Aoust et al. 2021).

The selection of an appropriate RNA isolation kit is a key component of processing samples that can have a major impact on the results yielded. This study aims to provide an overview of the efficacy and efficiency of four common RNA isolation kits produced by Zymo, Qiagen, and New England Biolabs when surveilling a small population for SARS-CoV-2 in wastewater. The endpoints examined in this study include a comparison of viral detection across all four kits as well as a qualitative description of each method.

## 2. Materials and Methods

### 2.1 Sample Collection

Wastewater was collected from 4 buildings in Waco, Texas on Baylor University’s campus. Sites A-C were collected from 3 dormitories and with a total, combined population of approximately 850 students at time of collection. Included in these sites was a dormitory that consistently yielded non-detectable values (site B). An additional isolation site, (Site I) was also included, this site housed an unknown number of students who were isolated due to active SARS-CoV-2 infections.

Composite 8.64-liters of untreated raw wastewater samples were collected over a 24-hr period in polypropylene bottles from 11:00 am on 10/6/2020 to 10:45 am on 10/7/2020. ISCO model 6712 automatic samplers were programmed to collect composite samples in 90 mL increments every 15 minutes. The sample bottle chamber was filled with ice to keep the samples cold. Upon collection, each composite sample was mixed by hand and an aliquot was poured into a 250 mL polypropylene centrifuge bottle (Fisherbrand, catalog # 14-375-352) and stored on ice. Samples were processed approximately 1 hour after collection.

### 2.2 RNA Concentration and Extraction

Following collection, raw wastewater samples were concentrated by centrifuging each 250 mL sample in the original bottle at 4°C for 45 minutes on a coast deceleration setting so as not to disturb the pellet (AVANTI JXN 26, JS-7.5 rotor, 4700 RCF). Supernatant (150 mL) was collected and aliquoted for use in three of the extraction kits. The remaining 100 mL pellet was resuspended and used for the pellet-based extraction kit.

The resulting 150 mL filtrate was further concentrated using ultrafiltration with AMICON 15 mL conical filtration tubes (Sigma Aldrich; UFC901024). Filtrate was loaded in increments of 15 mL until the total collection volume had been reduced to∼ 1000 µL (5000 RCF, Eppendorf A-4-62 swing-bucket rotor with adaptors). In some cases, the filters became clogged and a new ultrafilter was required and when necessary the sample was transferred to a new filtration tube. Concentrate was then aliquoted (∼250 µL) and RNA exactions carried out using extraction kits 1-3: Qiagen All Prep PowerViral DNA/RNA kit (80244), New England BioLabs Monarch RNA MiniPrep Kit (T2010S) and Zymo Quick RNA-Viral (R1034) following manufacturer instructions.

For the pellet, 45 out of the 100 mL of the resuspended pellet was collected into 50 mL conical tubes. Urine conditioning buffer (3.150 mL, D3061-1-140) was added thoroughly mixed, then centrifuged (5000 RCF x 15 min, RT; Eppendorf FA-45-6-30 fixed rotor). Following centrifugation, the remaining filtrate was removed (∼47.9 mL) leaving ∼ 250 µL concentrated pellet composite. To this, 750 µL of RNA/DNA shield (Zymo, R1100-250) was then added to the sample and held at 4°C until extraction. All extractions took place in an RNAse, DNAse-free hood environment, following manufacturers guidelines. A field blank using in-house tap water was run for primary concentration/processing integrity. RNA extraction blanks using nuclease free water were included with each extraction batch and kit.

### 2.3 PCR Analysis

Quantification of viral load was determined via RT-qPCR (QuantStudios 6 Flex) using New England BioLabs Enzyme and Probe Master kit (New England Biolabs, Catalog E3006X) with IDT N2 RUO primers (IDT, catalog #10006713). Each plate contained triplicates of each condition; whose CT’s were averaged to get the mean CT for each sample for each extraction method. The standards used contained a linearized 200,000 cp/uL N plasmid standard (IDT, catalog #10006625) which was diluted into a 10,000 copies/µL stock. Each plate’s standard curve was conducted using an 8 series dilution, starting with 10,000 copies/µL to 2.441 copies/µL. For the LOQ, CT values corresponding with a value of less than 2.441 copies/µL were listed as “not detected” values. In order to accurately assess whether inhibitors are acting upon collected samples, 1:2 dilutions were made from the RNA extract directly prior to plate analysis. Non-template controls confirmed PCR integrity. More information on PCR analysis can be found in the Supplemental Information.

## 3. Results

To benchmark the performance of the kits, the controls (viz. field blanks, extraction blanks and PCR blanks, and no template controls) were first all confirmed to be non-detectable for SARS-CoV-2. Thereafter, the individual sites were examined. Site B acts as a control for the methodology, with consistency low incidence levels historically recorded. For this sampling location, Site B resulted in a non-detectable sample across all RNA extraction kits evaluated (Table 1), with contrastingly high viral loads identified at the isolation site. In contrast, Site A and C both tested positive, with detectable levels consistent even between dilutions, with the exception of the Monarch RNA Miniprep Kit, which saw a non-detected value for the original sample, yet when diluted yielded a highly positive value for site A. For this specific kit, the lack of an inhibitor removal step may have allowed for a co-concentration of inhibitors to occur in this sample for this kit, where dilutions also diluted inhibitors, allowing for better detection. Otherwise, our samples performed better undiluted. This could be due to low viral loads in an already diluted matrix. There were differences between each of the kits’ amplifications, of which the Qiagen kit performed consistently between dilutions, potentially indicating adequate inhibitor removal, but yielded lower values compared to the other kits.

**Table 1:** Campus sites and SARS-CoV-2 detection in Copies/L with 4 different commercial RNA extraction kits.

| **Copies/Liter + Coefficient of Variation (CV)** |  |  |  |  |
| --- | --- | --- | --- | --- |
|  | **New England Biolabs Monarch RNA MiniPrep Kit** | **Qiagen AllPrep PowerViral DNA/RNA Kit** | **Zymo Quick RNA-Viral Fecal/Soil Microbe Microprep Kit** | **Zymo Quick RNA-Viral with Inhibitor Removal** |
| **Site A** |  |  |  |  |
| **undiluted** | Not detected | 6.2E+03<br>(19% CV) | 1.3E+04<br>(20% CV) | 3.9E+04<br>(11% CV) |
| **1:2** | 1.6E+04<br>(8% CV) | 5.3E+03<br>(38% CV) | 1.1E+04<br>(12% CV) | 4.0E+04<br>(16% CV) |
| **Site B** |  |  |  |  |
| **undiluted** | Not detected | Not detected | Not detected | Not detected |
| **1:2** | Not detected | Not detected | Not detected | Not detected |
| **Site C** |  |  |  |  |
| **undiluted** | 1.2E+04<br>(29% CV) | 5.7E+02<br>(87% CV) | 1.2E+04<br>(38% CV) | 1.0E+04<br>(21% CV) |
| **1:2** | 1.0E+04<br>(12% CV) | 2.5E+02<br>(74% CV) | 7.3E+03<br>(7% CV) | 9.8E+03<br>(10% CV) |
| **Site Isolation** |  |  |  |  |
| **undiluted** | 4.5E+07<br>(5% CV) | 2.1E+06<br>(17% CV) | 1.7E+07<br>(26% CV) | 2.3E+07<br>(8% CV) |
| **1:2** | 2.9E+07<br>(21% CV) | 1.2E+06<br>(5% CV) | 1.7E+07<br>(4% CV) | 1.6E+07<br>(9% CV) |

Between the kits, the Zymo Quick RNA-Viral with Inhibitor Removal consistently yielded the highest resulting viral loads. There were similarities between the Zymo Fecal Kit and the Quick-Viral RNA kit processes, including inhibitor removal steps. On average, the Zymo Fecal Kit values ranging from approximately 7,300 to 17,000,000 copies/L whereas the Zymo Quick-Viral RNA kit ranged from 9,800 to 23,000,000 copies/L. The Zymo Fecal Kit recovered approximately 73% (+/- 38%) more SARS-CoV-2 RNA than the Zymo Quick-Viral RNA kit per site. Examining the % Coefficients of variation between each site, each method, and each dilution factor was between 4%-87%. The Zymo Quick-Viral RNA kit had the lowest Coefficients of variations, whereas Qiagen’s kit had some of the higher values. The Zymo Fecal Kit pellet showed dilutions may improve Coefficients of variation values. CV values were calculated based off the final calculated cp/L. CV values to interpret PCR integrity can be found in Supplemental Table 4.

In regards to the isolation site, the New England BioLabs kit obtained the highest viral load at 45,000,000 copies/L in the original sample, and 28,000,000 copies/L in the diluted sample. However, New England BioLabs kit was the most variable in terms of consistency between dilutions. Here, the original sample carried a higher load (45.3%) than the diluted sample. The Zymo Fecal kit had the least variation between the dilution series (0.99%), as we saw with the site comparisons.

The impact of PCR inhibitors was examined using the isolation location (Site I), with little difference observed in the Zymo Fecal kit between concentrate and diluted sample in comparison to other kits. Interestingly, cp/L at the isolation site is markedly lower in the Qiagen kit compared to others which is surprisingly considering that this kit has been used heavily in wastewater testing of large municipalities (Fig. 1).

**Figure 1:**
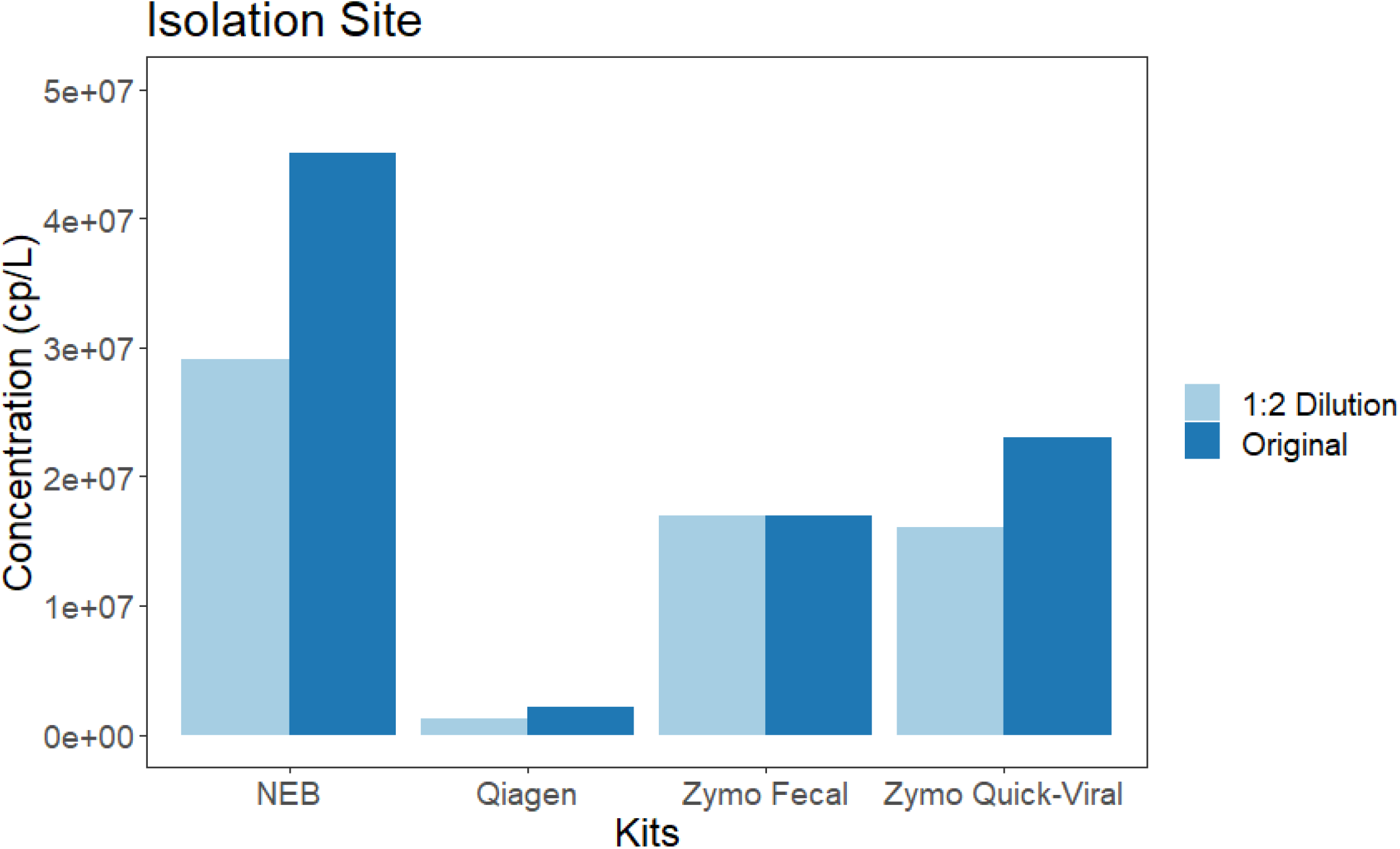
Investigating impacts of dilutions between the four kits for a known positive isolation site. Impact of PCR inhibitors were examined using the isolation location (Site I), with little difference observed in the Zymo Fecal kit between concentrate and diluted sample in comparison to other kits. Interestingly, cp/L at the isolation site is markedly lower in the Qiagen kit compared to others which is surprisingly considering that this kit has been used heavily in wastewater testing of large municipalities.

Impact of PCR inhibitors were examined using the isolation location (Site I), with little difference observed in the Zymo Fecal kit between concentrate and diluted sample in comparison to other kits. Interestingly, cp/L at the isolation site is markedly lower in the Qiagen kit compared to others which is surprisingly considering that this kit has been used heavily in wastewater testing of large municipalities.

### 3.2 Population Normalization

Normalizing the data by population is another key in determining hotspots for potential outbreaks. If the viral load per resident is high relative to the surrounding sites, this could be a potential indicator of an upcoming outbreak. Following normalizing to population size, Site A population (539) and Site C population (153), there is consistent differences in viral loads between extraction kits and between sites, with the Qiagen kit reporting markedly lower viral levels then the other kits examined. (Figure 2).

**Figure 2:**
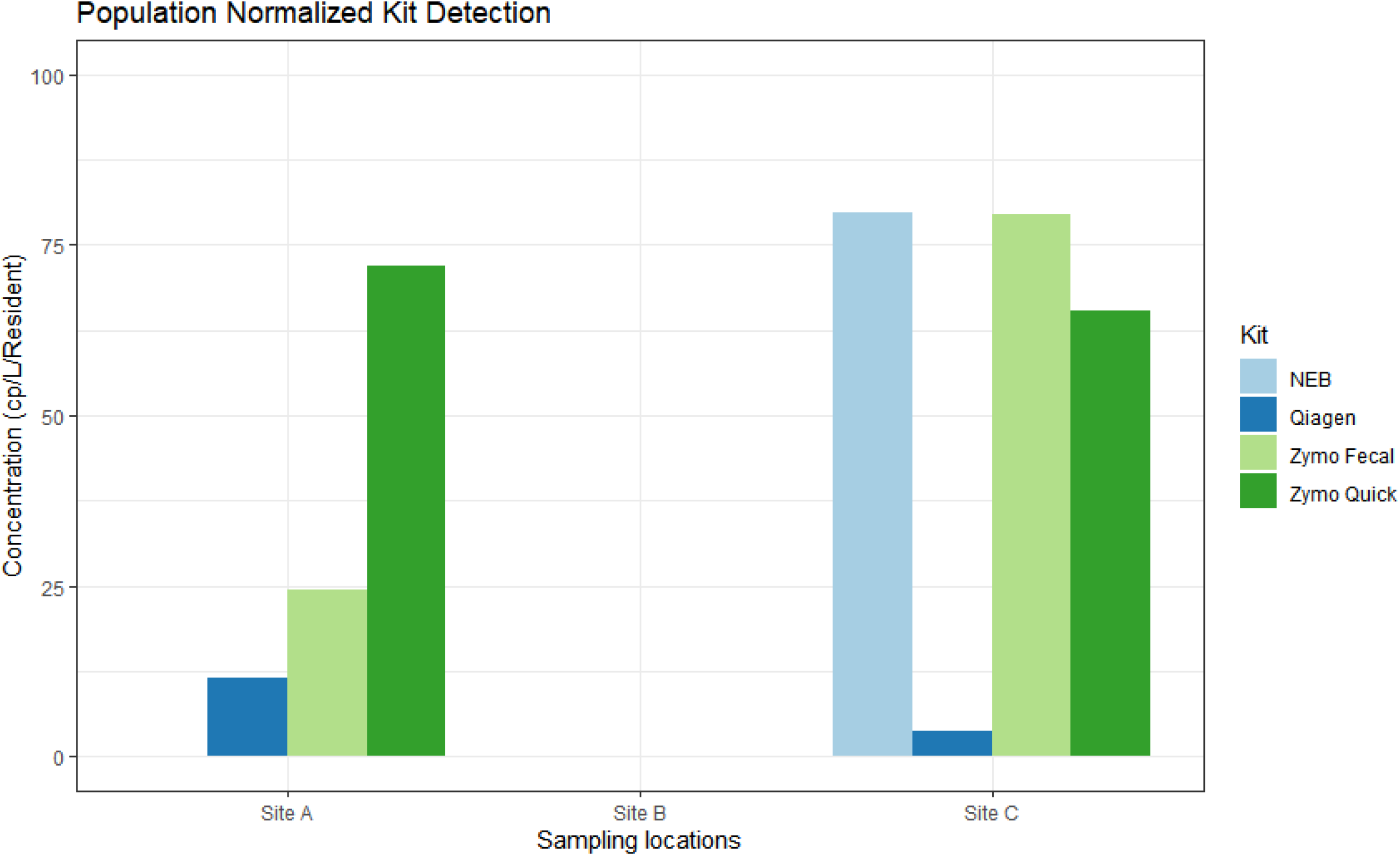
Average Copies/L normalized by population. Average Copies/L was calculated by dividing the number of residents for a specific site for the original, non-diluted sample.

### 3.3 Qualitative Results

Previous work has found that there is a lack of qualitative information for wastewater concentration and RNA extraction methodologies (Ahmed et. al 2020a). Therefore, we have provided qualitative information based on previous experiments. All times are approximate, and prices are listed as current for time of publication (Table 2). Footnotes can be found in Supplemental Information.

**Table 2:**
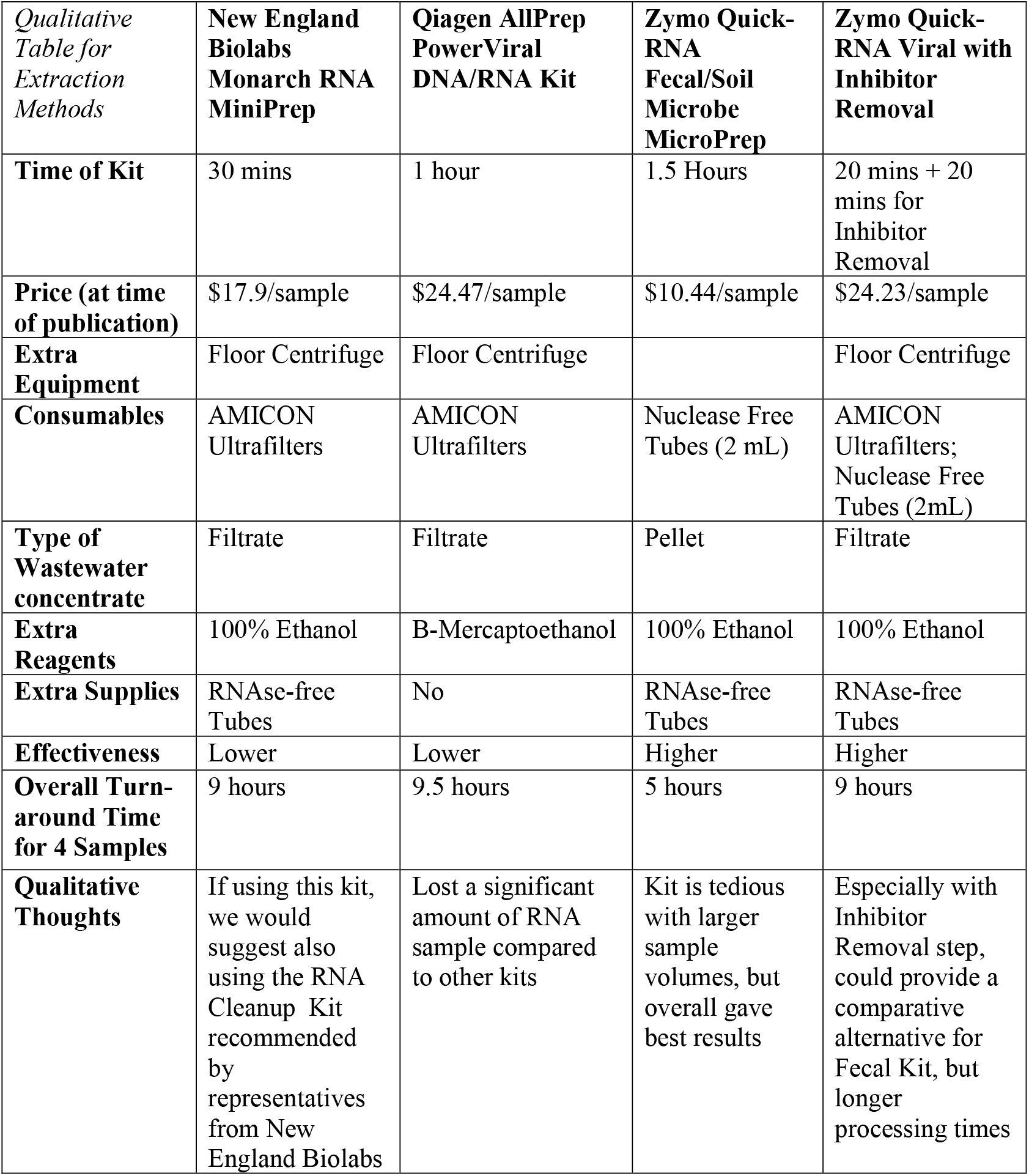
Qualitative descriptions for each RNA extraction kit used.

These times are total working hours and assume no breaks in between each section of processing, for only four samples. There is also the assumption that the technician has conducted the protocols prior to beginning the process. It should be noted that at the time of publication, the Zymo Quick-RNA Fecal/Soil Microbe MicroPrep Kit has been renamed as the Zymo Environ Water RNA Kit (Zymo Research, catalog #R2042), in which the only difference is that the urine conditioning buffer and DNA/RNA shield are included with the kit purchase.

Filtrate-based methods that use ultrafiltration considerably increased processing times due to long centrifuge runs and the potential for clogging filters (Table 2). The Zymo Quick-RNA Fecal/Soil MicroPrep kit required the most time, but yielded results more consistently across the board. This methodology also does not require the use of a floor centrifuge, meaning it may be available to a wider audience looking to start local surveillance.

## 4. Discussion

The ongoing pandemic of SARS-CoV-2 has allowed for greater expansion of wastewater based epidemiology (WBE) as a tool for public health surveillance. Recent testing conducted at the University of Arizona demonstrated the ability for campus wastewater surveillance to be used as an early detection system to pinpoint potential hot spots and isolate individuals, even before symptoms show (Betancourt et. al 2021). Historically, WBE has focused mostly on enteric viruses, where many of these viruses are non-enveloped. Since SARS-CoV-2 is an enveloped virus, this provides additional challenges for its concentration and detection in wastewater (Ye et. al 2016, Polo et. al 2020, Torii et. al 2021). Traditional methods of RNA extraction, such as polyethylene glycol precipitation (PEG) are not as effective at concentrating enveloped viruses since the lipid membrane is more sensitive and possibly degraded by organic solvents like chloroform (Polo et. al 2020) and in past publications for SARS-CoV-2 have returned lower recovery efficiencies (Ahmed et. al 2020b, Torii et. al 2021). The current study populations varied from 140-540 persons per site, compared with upwards of 10,000 or more for a municipal wastewater treatment plant. Since PEG precipitations require small initial volumes, low concentration/high volume samples are at a disadvantage and rely heavily on primary concentration methods to be effective (Lu et. al 2020). The Qiagen AllPrep PowerViral DNA/RNA kit has been used frequently in testing municipal wastewater for SARS-CoV-2 RNA, where a higher population density is contributing to the collection sample. When considering smaller populations and thus smaller RNA inputs, the Qiagen kit did not perform as well in our study. This could be due to the selectivity of combined RNA/DNA columns, or the lack of DNA/RNA preservation (such as DNA/RNA shield or urine conditioning buffer) throughout the long filtration steps.

Primary concentration from raw wastewater samples has been demonstrated as an obstacle in effectively concentrating viral loads (Hamouda et. al 2021, Ahmed et. al 2020b). Pellet-based methods have gained traction recently for higher quality and higher quantity of viral copies per sample (Kitamura et. al 2021, Pérez-Cataluña et. al 2021, D’Aoust et. al 2020, Graham et. al 2021). The filtrate based kits used in this study required larger sample volumes compared to the pellet-based kits, and the filtrate based kits also require an additional ultrafiltering step. Ultrafilter clogging was an issue with ultrafiltering, and loss of viral load is assumed with each tube transfer. Consumables associated with primary concentration, such as ultrafilters and 50 mL Falcon tubes, are becoming increasingly difficult to obtain due to backordering.

Finding effective methods to accurately identify low SARS-CoV-2 viral loads in wastewater are important for sectors such as hospitals, nursing homes, or schools, which have a lower flow rates than municipal wastewater treatment plants that are normally investigated during broad surveillances. The Qiagen All Prep PowerViral kit columns select for both DNA and RNA, where RNA could be potentially lost. New England BioLabs suggests the use of their RNA Cleanup Kit to aide in better recover/prevent RNA degradation. The Zymo Quick-RNA Fecal/Soil MicroPrep Kit included two forms of RNA preservation (DNA/RNA shield and urine conditioning buffer) which may have aided in its higher recoveries. Based on these results, we would recommend choosing a kit that specifically targets RNA extraction, and not a combined kit. This would allow for DNA removal as well, with which we saw increased detection in RNA viral loads. Homogenization of samples prior to extraction can aide in breaking viral capsids and releasing viral RNA. The Qiagen kit did not include filtrate homogenization steps, which could have affected its lower detection values. A kit that includes an inhibitor removal component is also vital, even in samples with low population density. The Monarch Kit did not include an inhibitor removal step during the extraction process, whereas the other kits involved at least one RNA inhibitor removal step. This would indicate that inhibitor removal was indeed necessary to see higher amplifications in CT values, since diluting inhibited samples is known to increase the efficiency of primer binding to cDNA, where inhibitors themselves are then diluted (Hata et. Al 2015) Dilutions could be conducted in the instance that a kit with inhibitor removal is unavailable.

A greater need for surveillance raises questions with cost and accessibility. For smaller sampling canvases, such as hospitals or nursing homes, a kit that can quantify smaller amount of the virus accurately is important. Further, the ability to conduct in house processing would greatly alleviate the expenditures of sending samples out to another lab. Time is also an important factor to consider while choosing an extraction method. Depending on the number of samples, as well as sample composition, time can vary greatly.

Since viral shedding through feces can be seen in both symptomatic and asymptomatic/presymptomatic patients, health officials can see larger trends in community prevalence than through nasal swabbing alone (Hart et. al 2020, Mizumoto et. al 2020, Treibel et. al 2020). Duration of viral shedding in symptomatic patients can vary anywhere from a matter 14-21 days (Wu, 2021). It is indicated that viral shedding through feces precedes symptoms of COVID-19, and thus is imperative that precautionary measures be taken as soon as possible to prevent widespread contagion throughout the sample population, even in low prevalence areas (Randazzo et. al 2020). Results from this study were used to choose a concentration method and RNA extraction kit for Baylor University’s campus wide wastewater surveillance program for the spring of 2021.

## 5. Conclusion

The results of this study show that a pellet-based RNA extraction kit that includes an inhibitor removal and RNA preservation step may yield the most consistent, timely, and accurate results. These additional steps may be why the Zymo Quick-RNA Fecal/Soil Microbe MicroPrep (also called the Zymo Environ Water RNA kit) was the most effective and efficient kit with the samples we used in the study. In contrast, the least effective kit was the Qiagen All Prep PowerViral DNA/RNA Kit. This was likely due to the absences of a preservation step and inefficiencies stemming from the kit selecting for both DNA and RNA. For filtrate based methods, we recommend using the Zymo Quick-RNA Viral kit as an effective and efficient method of wastewater based epidemiological analysis in concentrated wastewater samples, especially in smaller population densities.

## Data Availability

Data is available upon request

## Acknowledgements

We would like to acknowledge the financial support from Baylor University and for support from Dr. Bryan Brooks and Dr. Kevin Chambliss. We would also like to thank the Molecular Biosciences Center for the use of the equipment in the core and other reagents used in this study.

