## Supplemental information for "A comparison of four commercially available RNA extraction kits for wastewater surveillance of SARS-CoV-2 in a college population"

Dr. Joaquin N. Lugo

Department of Psychology and Neuroscience

One Bear Place 97334

Baylor University

Waco, TX 76798

**PCR Specifications**

Sample Preparation

Eluted RNA volumes ranged from 35 µL (Zymo) or 50 µL (Qiagen & New England Biolabs) respectively. Template (RNA; 5 µL) were added to each well and 15 µL of master mix was added.

Primer, Probe, RT sequences

The master mix was created by using the following calculations. The master mix was created using the calculations below **n* of samples.

**Table S1: Sample Master Mix**

| Luna Universal One-Step Reaction Mix | 10 µL |
| --- | --- |
| Luna Warmstart RT Enzyme Mix | 1 µL |
| IDT RUO N2 Primer | 1.5 µL |
| Nuclease Free Water | 2.5 µL |
| Extracted RNA | 5 µL |

Standard Linearization and Dilutions:

Plasmid standards (250 µL; stock 200,000 cp/µL; 2019-nCoV_N_Positive Control, IDT) were linearized using the New England Biolabs ScaI-HF (Cat. No. R3122S) restriction enzymes and diluted to 100,000 cp/µL with nuclease free water. Stocks of 10,000 cp/uL were created, aliquoted, and used fresh for each PCR reaction.

Serial dilutions ranging from 10,000 cp/µL to 2.44 copies/µL were utilized for standard curve. Standards were prepped on the benchtop to prevent plasmid contamination in the hood.

The master mix for the standards was created using the same kit for the samples, but with the following calculations:

**Table S2: Standards Master Mix**

| Luna Universal One-Step Reaction Mix | 10 µL |
| --- | --- |
| IDT RUO N2 Primer | 1.5 µL |
| Nuclease Free water | 7.5 µL |
| Standard DNA | 1 µL |

Instrumentation Used and Cycling Parameters

The Quantstudio Flex 6 system was used with Applied Biosystems software for thermocycling using the TaqMan reagent option with the following parameters:

**Table S3: Thermocycling Parameters**

| **Reaction Cycling Step** | **Temperature** | **Time** |
| --- | --- | --- |
| Initial RT Transcription | 55˚ C | 2 mins |
| Initial Denaturation | 95˚ C | 10 mins |
| Denaturation | 95˚ C | 15 seconds |
| Extension | 60˚ C | 1 min |

Calculations

Calculations were conducted using the slope of the linear standard curve.

Slope = CT/log_10_(std)

Amplification efficiency (EAMP)= 10^(-1/SLOPE)

All variations were analyzed in triplicates, with the CT values for each variation being averaged prior to further analysis. Any value with a CT higher than 39 was counted as a negative value. Copy number was calculated based on the following formula and dilution corrected:

COPIES/RXN (cp/rxn) = EAMP^(Y INTERCEPT- SAMPLE CT AVERAGE)^

Copies/L was determined using the following equation where:

Copies/L =	1000 *	((Copies/RXN)/ RNA template volume)* Total eluted RNA volume

Original sample volume (mL)

Where total eluted RNA volume is kit dependent.

From the total copy number, the total copies/L was deduced by assuming a total concentration of RNA from the original sample (i.e, total copy number in 35 µL is representative of the whole 45 mL sample)

Coefficient of Variation Values

CV percent values were calculated off final calculated cp/L values. This was to show differences in the final copy numbers that could be calculated for each sample. This was not to interpret PCR integrity. Below are the %CV values for the PCR CT triplicates.

Table S4: %CV values based off CT values

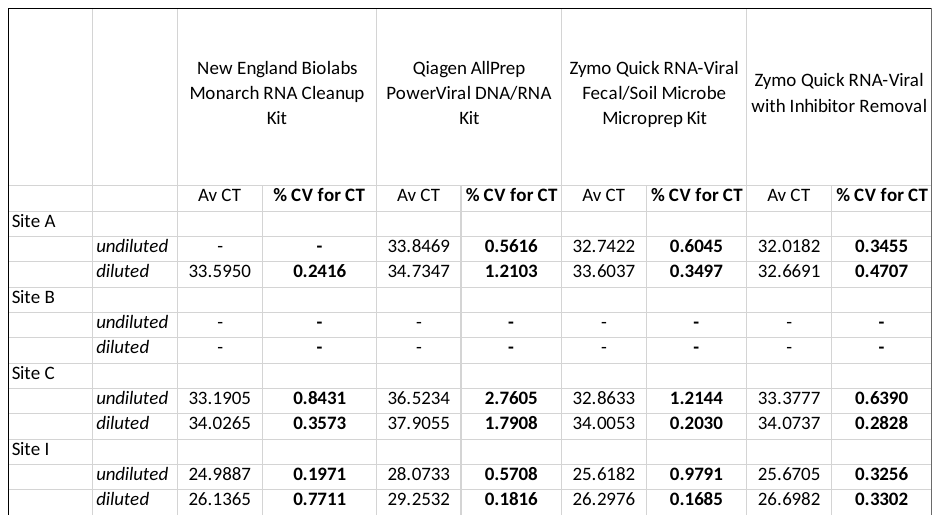
